# Discovery of tumour indicating morphological changes in benign prostate biopsies through AI

**DOI:** 10.1101/2024.06.18.24309064

**Authors:** Eduard Chelebian, Christophe Avenel, Helena Järemo, Pernilla Andersson, Anders Bergh, Carolina Wählby

## Abstract

**Background and Objective:** Diagnostic needle biopsies that miss clinically significant prostate cancers (PCa) likely sample benign tissue adjacent to cancer. Such samples may contain changes indicating the presence of cancer elsewhere in the organ. Our goal is to evaluate if artificial intelligence (AI) can identify morphological characteristics in benign biopsies of men with raised PSA that predict the future detection of clinically significant PCa during a 30-month follow-up.

**Methods:** A retrospective cohort of 232 patients with raised PSA and benign needle biopsies, paired by age, year of diagnosis and PSA levels was collected. Half were diagnosed with PCa within 30 months, while the other half remained cancer-free for at least eight years. AI model performance was assessed using the area under the receiver operating characteristic curve (AUC) and attention maps were used to visualise the morphological patterns relevant for cancer diagnosis as captured by the model.

**Key findings and Limitations:** The AI model could identify patients that were later diagnosed with PCa from their initial benign biopsies with an AUC of 0.82. Distinctive morphological patterns, such as altered stromal collagen and changes in glandular epithelial cell composition, were revealed.

**Conclusions and Clinical Implications:** AI applied to standard haematoxylin-eosin sections identifies patients initially diagnosed as negative but later found to have clinically significant PCa. Morphological patterns offer insights into the long-ranging effects of PCa in the benign parts of the tumour-bearing organ.

**Patient Summary:** Using AI, we identified subtle changes in normal prostate tissue suggesting the presence of tumours elsewhere in the prostate. This could aid in the early identification of potentially high-risk tumours, limiting overuse of prostate biopsies.

## 1. Introduction

Prostate cancer (PCa) remains a significant health concern worldwide, affecting 1.4 million men annually [1]. While most cases follow a rather indolent course, some progress to aggressive tumours resulting in fatal outcomes [2]. The current diagnostic paradigm relies on raised prostate-specific antigen (PSA) in blood samples, followed by magnetic resonance imaging (MRI) and prostate needle biopsies [3]. However, due to the low specificity of PSA tests, false negative MRI and the limited coverage of needle biopsies, these may fail to detect PCa even when an aggressive tumour is present [4, 5, 6, 7]. Consequently, there is a need for novel diagnostic and prognostic markers capable of identifying early changes associated with the presence of a clinically significant PCa. Such changes could be detected in apparently histologically normal tissue adjacent to tumours. Multiple studies have shown that the benign parts of a tumour-bearing organ and a non-tumour-bearing organ are different [8, 9, 10]. A common explanation to this is the cancer field hypothesis which states that main parts of an organ are affected by carcinogenic agents resulting in precancerous genetic changes in the epithelial compartment [11]. Alternatively, to grow and spread, tumours need to instruct surrounding normal tissue to assist. Bergh and colleagues demonstrated alterations in the epithelial and stroma compartments of the tumour-bearing prostate in fully immune-competent rats, with the nature and magnitude of the effects being associated with tumour proximity, aggressiveness and metastatic capacity [9, 12, 13, 14, 15, 16, 17]. Studies in patients have confirmed similar changes and related them to patient outcome [18, 19, 20, 21, 22]. We have termed these changes as tumour instructed (and thus indicating) normal tissue (TINT) and proposed that they could be used as novel diagnostic and prognostic markers [9].

Recognising that prostate needle biopsies may fail to sample tumour tissue even when cancer is present, we conducted a retrospective analysis using a dataset of patients with raised PSA and whose initial biopsies only contained benign prostate tissue. We paired patients who, within 30 months after an initial benign diagnosis, were diagnosed with PCa of varying histological grades (ISUP grades) with patients who remained cancer-free for at least eight years after their initial benign diagnosis, matching them for age and PSA levels. Deep learning techniques, specifically weakly-supervised learning [23], allow to handle the challenge of having only patient-level information, in this case the cancer grade detected in a patient at subsequent biopsy rounds. This approach enabled the analysis and comparison of the morphological changes in hundreds of patients, a task unattainable through visual inspection alone. Our hypothesis is that it is possible to identify high-risk patients that were later diagnosed with clinically significant PCa by examining their initial benign biopsies. Additionally, we analysed the histopathological features responsible for the model performance, contributing to the understanding of the morphological changes potentially induced in TINT.

## 2. Patients and Methods

### 2.1. Study design

We collected a retrospective cohort of 232 patients from the University Hospital of Umeå (*Norrlands universitetssjukhus*) in Umeå, Sweden over the period spanning from 1997 to 2016. Ethical approval was granted from *Regionala etikprövningsnämnden i Umeå*, DNR 2010/366-31M. The study focuses on men (age 65 ± 7) with raised serum PSA (11.8 ± 7.5), an initial benign diagnosis in prostate needle biopsies (excluding cases with precancerous lesions such as PIN) and with subsequent separation in diagnostic trajectories. While some patients were finally diagnosed with PCa with varying ISUP grades at subsequent biopsy rounds within 30 months, others, despite subsequent re-biopsy rounds, remained cancer-free for a period of at least eight years. The patients were paired based on similar age, year of diagnosis and PSA level upon initial diagnosis.

As patients diagnosed with ISUP grade 1 typically receive recommendations for active surveillance [3, 24], the primary objective of this study is to find differences in the initial diagnostic morphological characteristics between (1) patients who either remained cancer-free or were diagnosed with ISUP grade 1, termed low-risk, and (2) patients who were diagnosed with clinically significant cancer, that is with ISUP grades greater than 1 (ISUP*>*1), termed high-risk.

### 2.2. Image acquisition

The patients underwent standard of care needle biopsy examinations to determine their benign state. For each patient, 10–12 needle biopsies of approximately 10 mm in length were extracted at prespecified locations. We digitised the available slides using the 3D Histech Pannoramic 250 scanner (3D HISTECH Ltd., Budapest, Hungary) at a magnification of 20× and a pixel size of 0.2428 μm/pixel

### 2.3. Deep learning workflow

In the field of artificial intelligence, problems where the available labels are image-level, as opposed to pixel-level annotations, are termed weakly-supervised problems. Our study can be framed within this paradigm, as our knowledge is limited to whether a specific slide originates from a patient who either was later diagnosed with PCa or remained benign after initial benign diagnosis. That is, the initial biopsy which we analyse showed no suspicious findings in different rounds for the pathologists and we use the future status as a label. This approach enables the automated analysis and comparison of morphological characteristics across hundreds of slides, facilitating the exploration of the subtle differences that guide a model’s decision.

We employed a customised version of the state-of-the-art clustering constrained attention multiple-instance learning (CLAM) algorithm [23]. Multiple-instance learning works under the assumption that negative slides — originating from patients who continued to be benign — are negative evidence, while positive slides — originating from patients who were later diagnosed with PCa — contain at least one region of the tissue that contributes as positive evidence. These regions in positive slides are then analysed in pursuit of TINT changes.

The initial step of our workflow includes detection and segmentation of tissue within digitised biopsies, which are subsequently divided into non-overlapping tiles of 256 × 256 pixels or approximately 62 × 62 *μm*. From each tile, representations are extracted employing self-supervised learning [25], enabling the learning of meaningful image representations without the requirement for manual annotations. With each slide represented as a collection of tile feature vectors corresponding to these representations, we train the weakly-supervised model using the slide-level labels. A complete description of the workflow is available in Appendix A.

### 2.4. Statistical analysis

The patient cohort was partitioned into a development cohort, constituting 80% of the subjects for model optimisation, using 5-fold cross validation. To assess generalisation of the best performing model on validation, we used the remaining 20% of subjects as independent test set. We ensured that slides from the same patient did not appear in different cohorts and we balance the cohorts in terms of ISUP grade, PSA level and age distribution.

The performance of the model was evaluated via the area under the receiver operating characteristic curve (AUC), assessed both at slide-level and patient-level. Patient-level predictions were obtained by considering a patient high-risk if at least one of their slides was classified as high-risk.

To interpret the morphological characteristics driving the classification process, we employed uniform manifold approximation and projection for dimension reduction (UMAP) [26] on the the tile-level representations. This approach enabled the identification of tumour associated changes within the initial benign biopsies by visualising the representations in a two-dimensional space.

## 3. Results

### 3.1. Baseline characteristics

The final cohort included 213 patients with 587 biopsy slides as detailed in Figure 1. 19 patients were excluded due to sample reasons, either for not being H&E-stained prostate needle biopsies or for missing data. No further patients were excluded, although six additional slides were discarded due to imaging reasons. We publicly share the final cohort in [27].

**Figure 1:**
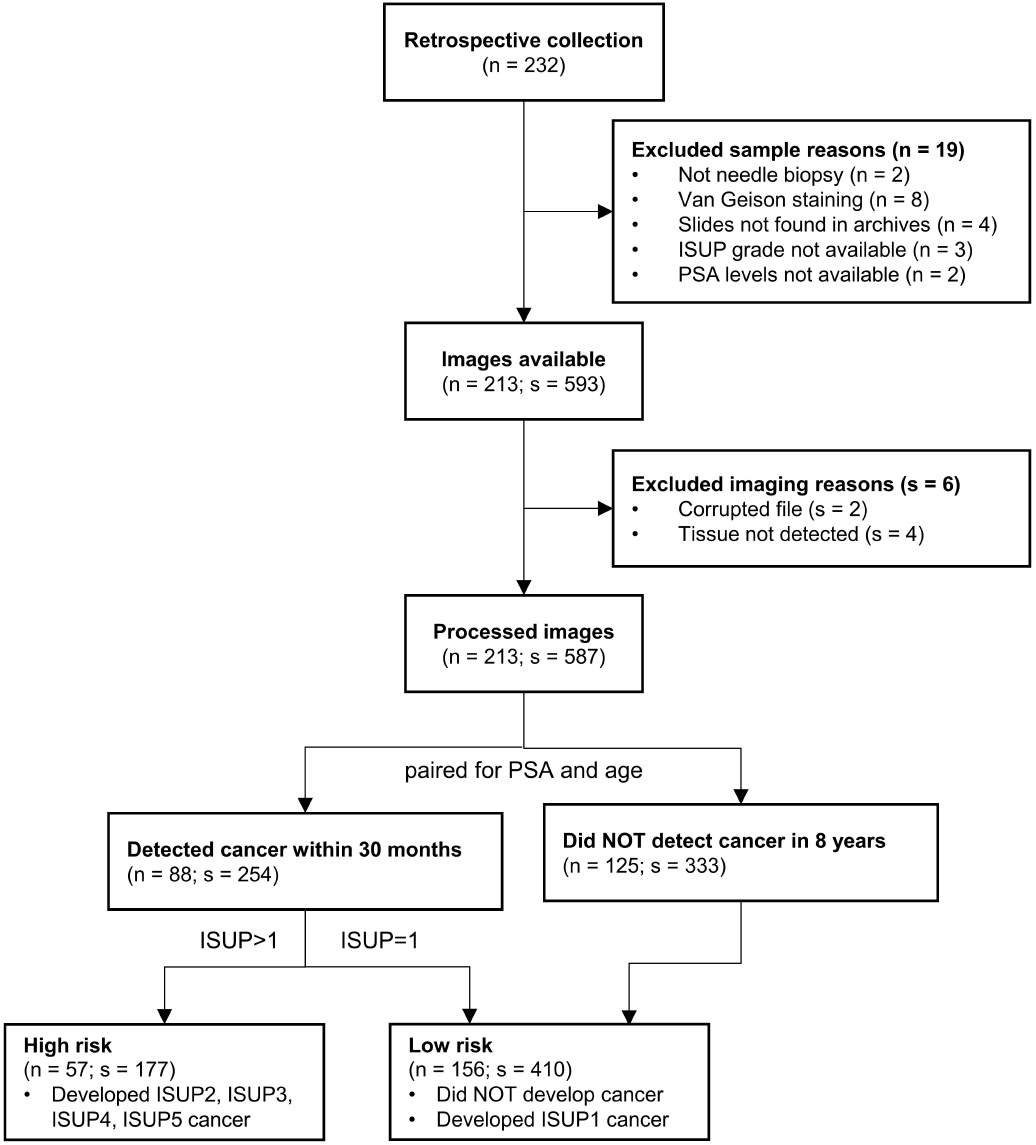
Consort diagram (n=patients; s=slides)

Table 1 summarises the characteristics of the final cohort, and the partitions into the model development cohort and the test cohort.

**Table 1:** Baseline characteristics. Percentages compared to the complete cohort per column.

|  | Total<br>(n=213) | Development<br>(n=170) | Test<br>(n=43) |
| --- | --- | --- | --- |
| **Age (years)** |  |  |  |
| < 50 | 1 (0.5%) | 1 (0.6%) | 0 (0.0%) |
| 50–55 | 14 (6.6%) | 12 (7.1%) | 2 (4.7%) |
| 55–60 | 36 (16.9%) | 30 (17.6%) | 6 (14.0%) |
| 60–65 | 55 (25.8%) | 46 (27.1%) | 9 (20.9%) |
| 65–70 | 55 (25.8%) | 40 (23.5%) | 15 (34.9%) |
| 70–75 | 33 (15.5%) | 24 (14.1%) | 9 (20.9%) |
| > 75 | 19 (8.9%) | 17 (10.0%) | 2 (4.7%) |
| **PSA (ng/mL)** |  |  |  |
| < 3 | 1 (0.5%) | 1 (0.6%) | 0 (0.0%) |
| 3–5 | 21 (9.9%) | 17 (10.0%) | 4 (9.3%) |
| 5–10 | 97 (45.5%) | 76 (44.7%) | 21 (48.8%) |
| > 10 | 94 (44.1%) | 76 (44.7%) | 18 (41.9%) |
| **ISUP grade** |  |  |  |
| Benign | 125 (58.7%) | 101 (59.4%) | 24 (55.8%) |
| ISUP 1 | 31 (14.6%) | 24 (14.1%) | 7 (16.3%) |
| ISUP 2 | 11 (5.2%) | 10 (5.9%) | 1 (2.3%) |
| ISUP 3 | 19 (8.9%) | 14 (8.2%) | 5 (11.6%) |
| ISUP 4 | 19 (8.9%) | 14 (8.2%) | 5 (11.6%) |
| ISUP 5 | 8 (3.8%) | 7 (4.1%) | 1 (2.3%) |

### 3.2. Model performance

Figure 2 shows the model performance results. We identified slides coming from patients who would be diagnosed with a clinically significant PCa (ISUP2–5) with an *AUC* = 0.81, and sensitivity of 0.81 at false positive rate of 0.26 on the test set. At patient level, we achieved an *AUC* = 0.82 with a sensitivity of 0.92 at false positive rate of 0.32. Considering these patients were not identified at all upon initial diagnosis, we consider the sensitivity compensates for the false positive rates.

**Figure 2:**
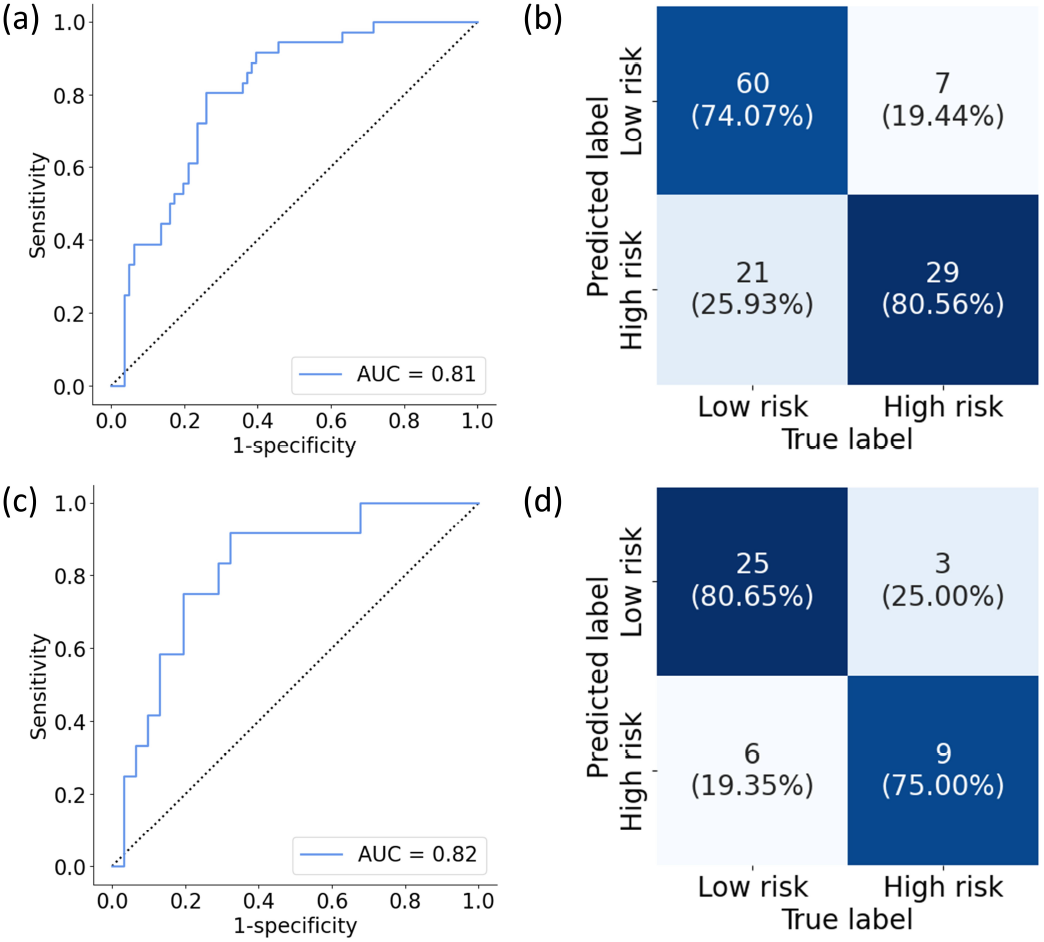
Model performance on the test set. Per slide (a) ROC curve and (b) contingency matrix. Per patient (c) ROC curve and (d) contingency matrix.

Figure 2b and Figure 2d show the confusion matrices predicted patients into low-risk (cancer-free and ISUP1) and high risk (ISUP2–5). A more detailed view dividing the patients into their true ISUP class is shown in Appendix B. Interestingly, patients who developed ISUP1 were easier to identify as low risk than patients who continued being benign. There was also a difference of performance in identifying patients who would develop high grade PCa (ISUP4–5) and patient at intermediate risk such as ISUP 2 or ISUP 3.

### 3.3. Morphological interpretation

Figure 3 shows the UMAP space of the most and least attended tiles from the correctly classified slides in the test set.

**Figure 3:**
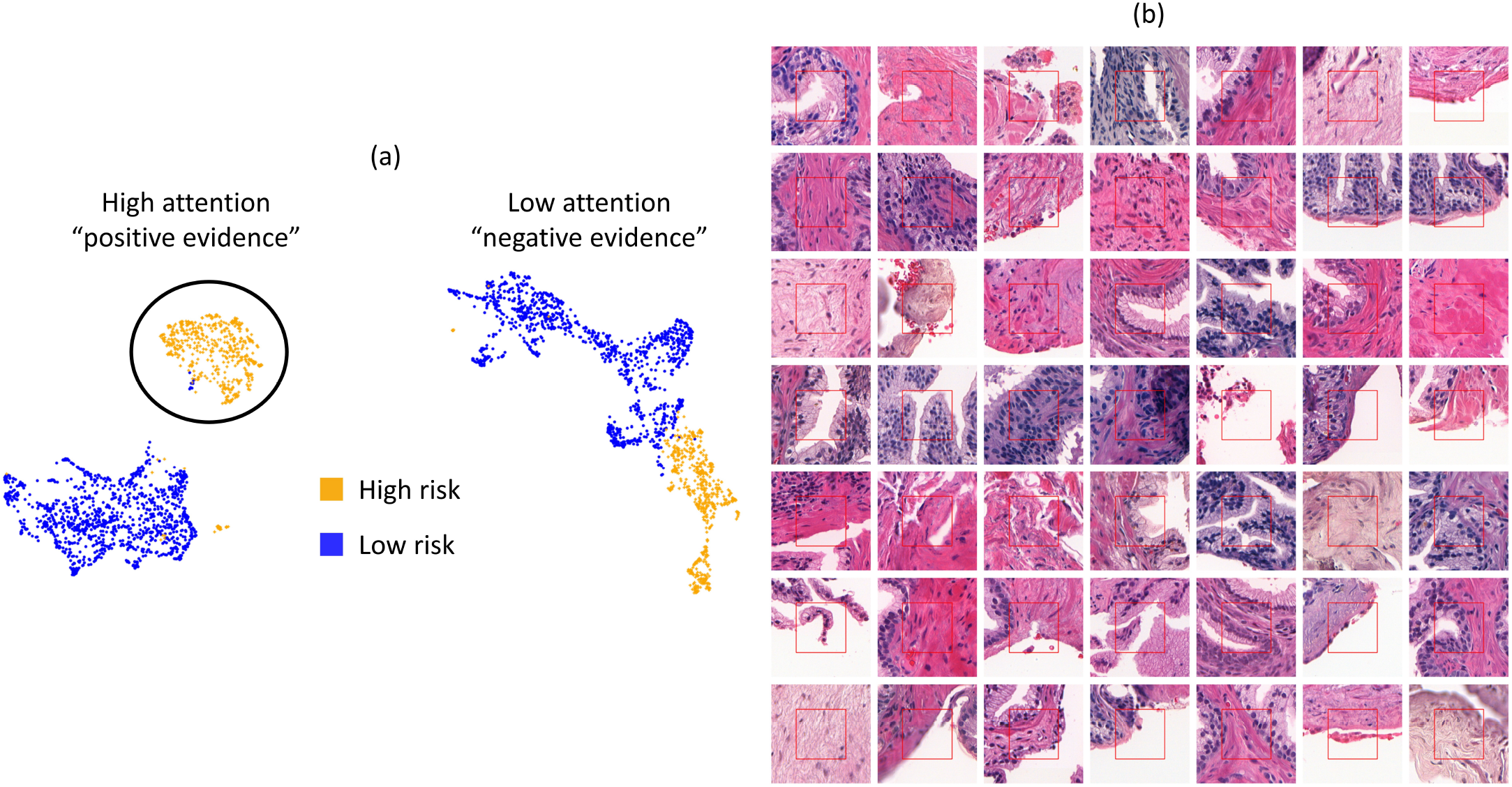
(a) UMAP space of the 20 most and least attended tiles from the correctly classified slides in the test set. (b) Sample of the most attended morphologies that drove the classification, the *positive evidence* for being high risk. The red square in the middle is the actual tile size, the rest is shown for context.

Tiles naturally clustered between positive and negative evidence. While low attended patches are more tightly packed regardless if they are low or high risk, highly attended tiles clearly clustered between low and high risk patients. In Appendix C we show the representation space for the whole test cohort.

An interactive overlay of attention regions is shared at https://prostate-attention.serve.scilifelab.se/. Note that these attention heatmaps are smoothed for better visualisation. We present the initial negative needle biopsy for two patients that were eventually diagnosed with ISUP3 and ISUP4 and the model was able to recognise them.

The sample of the morphology that drove the classification shows that changes within the tissue stroma compartment were common and informative and apparently characterised by increased collagen and reduced numbers of smooth muscle cells. Epithelial characteristics could also drive classification, but the epithelial phenotype involved is not evident at this stage.

## 4. Discussion

The present study suggests that AI, applied to routine H&E-stained needle biopsies, can identify morphological changes in men with elevated PSA, distinguishing between those likely to be diagnosed with significant PCa within 30 months of follow-up and those expected to remain low-risk for eight years with a AUC of 0.82. Previously, AI methods have been successfully used to detect prostate cancer in needle biopsy cores [28] to reduce pathologist work burden by trying to replicate the diagnostic process. More recently, similar techniques have been able to differentiate tumour-adjacent benign prostate tissue in the same biopsy from prostate tissue in tumour-free men [29]. We went a step further, leveraging novel weakly-supervised learning techniques to analyse the initial benign biopsies of patients that were later diagnosed with PCa. Weakly-supervised methods have also been successful for large-scale screening in other diseases [30].

Multiple studies have demonstrated that prostate tumours in patients are associated with changes in the benign parts of the organ [8, 9, 10]. Those changes have been detected utilising various methods, including alterations in transcriptome [10, 31, 32], metabolome [33, 34], morphology [8, 18, 19, 21, 22, 16, 17] or epigenetic marker expression [35, 36]. Such alterations could be used to diagnose and prognosticate PCa indirectly by examining the adjacent benign parts of the prostate [9]. Specifically, they could be used to evaluate the need of re-biopsy in cases with raised PSA and initially negative prostate biopsies [9, 37, 36]. Additionally, previous studies in experimental models [12, 13, 15, 38] and in patient samples [19, 21, 22, 14, 17] indicate that the magnitude and nature of the changes detected in the benign parts of the tumour-bearing prostate are related to tumour aggressiveness, proximity and patient outcome. For instance, the magnitude of some morphological changes, such as the increased vascular density and accumulation of macrophages adjacent to a tumour, were related to the tumour ISUP grade. Changes in benign tissue adjacent to ISUP1 tumours were more discrete while changes around ISUP 4–5 were more pronounced [17].

Considering this background, we hypothesised that our model could differentiate changes in the benign parts of the prostate related to tumour grade, and particularly to identify high-risk cases in need of early re-biopsy. ISUP1 tumours are generally indolent so usually there is no need of early diagnosis and active treatment [24]. Importantly, our model could identify such cases and notably an ISUP1 tumour was by far the most common type detected among the cases with raised PSA later diagnosed with PCa. Some ISUP2 are candidates for active treatment while others are not and ISUP 3–5 tumours generally need early diagnosis and active therapy [3].

Our model could identify cases that were later diagnosed with ISUP 2–5 with high sensitivity, although it was less effective for ISUP4–5 cases. This could be due to many ISUP4–5 tumours showing high cell proliferation and rapid growth [17] so they may have been too small and too far away from the biopsies to have a significant effect on the initial biopsies. Some cases that remained cancer-free were grouped among high-risk cases. As tumour-adjacent benign tissue is characterised by biological processes such as inflammation and wounding response [13, 15, 38, 10, 17], inflammation or prostatitis could induce changes that could be misinterpreted as cancer-induced alterations by our model. It should be noted that the number of cases that were later diagnosed with high-grade tumours was rather low and further studies using larger cohorts are needed for more firm conclusions.

Our model was able to identify areas within biopsies that were particularly informative to identify cases that were later diagnosed with PCa. The exact nature of these changes needs to be clarified using, for example, spatial transcriptomics or immunohistochemisty of selected key markers. However, already at this stage, it can be concluded that informative changes are identified in discrete parts of the stroma and the glandular epithelium. In informative stroma areas reduced fraction of smooth muscle cells and increased collagen/altered matrix were observed. Interestingly, this highly resembles the development of reactive stroma within prostate tumours [39] and the signals causing a reactive tumour stroma may extend out into the surrounding tumour-bearing organ [9]. Importantly, as our AI achieved high sensitivity, it could be combined with tests for serum PSA variants [40] that have high negative predictive value.

## 5. Conclusions

In conclusion, AI was able to identify, from their initially benign H&E-stained needle biopsies, patients who were later diagnosed with clinically significant PCa. The discovered morphological characteristics can provide insights on the biological mechanisms of tumour indicating normal tissue in the tumour-bearing prostate. If these findings were confirmed, the methodology could be used as a complementary screening step.

## Data Availability

All data produced in the present study are available upon reasonable request to the authors

