## Appendix for "Discovery of tumour indicating morphological changes in benign prostate biopsies through AI"

### Appendix A. Deep learning workflow

The general workflow is based largely on the clustering constrained attention multiple instance learning (CLAM) model presented by Lu *et al.* [1]. The first step requires to detect the tissue from the background. This is done with the classical Ostu's thesholding technique on a smoothed version of the saturation channel of the HSV converted image. After morphological closing, an area filter is applied to discard small detected objects. The detected tissue is then divided into $256 \times 256$ patches at the highest resolution at $20\times$ magnification. An important difference with the standard workflow is that we extract features from the patches with a ResNet18 trained in a self-supervised way on 57 histopathological datasets [2]. This differs from the ResNet50 used by default which is trained on the natural image ImageNet dataset. This result in each $256 \times 256$ patch being embedded into a 512-dimensional feature vector. Finally, we train the model with the single-branch "small" version of the CLAM model for 100 epochs with early stopping and cross-entropy bag loss.

The implementation of the CLAM model can be found here: `https://github.com/mahmoodlab/CLAM` and the self-supervised model we used is shared here: `https://github.com/ozanciga/self-supervised-histopathology`.

### Appendix B. Contingency matrix

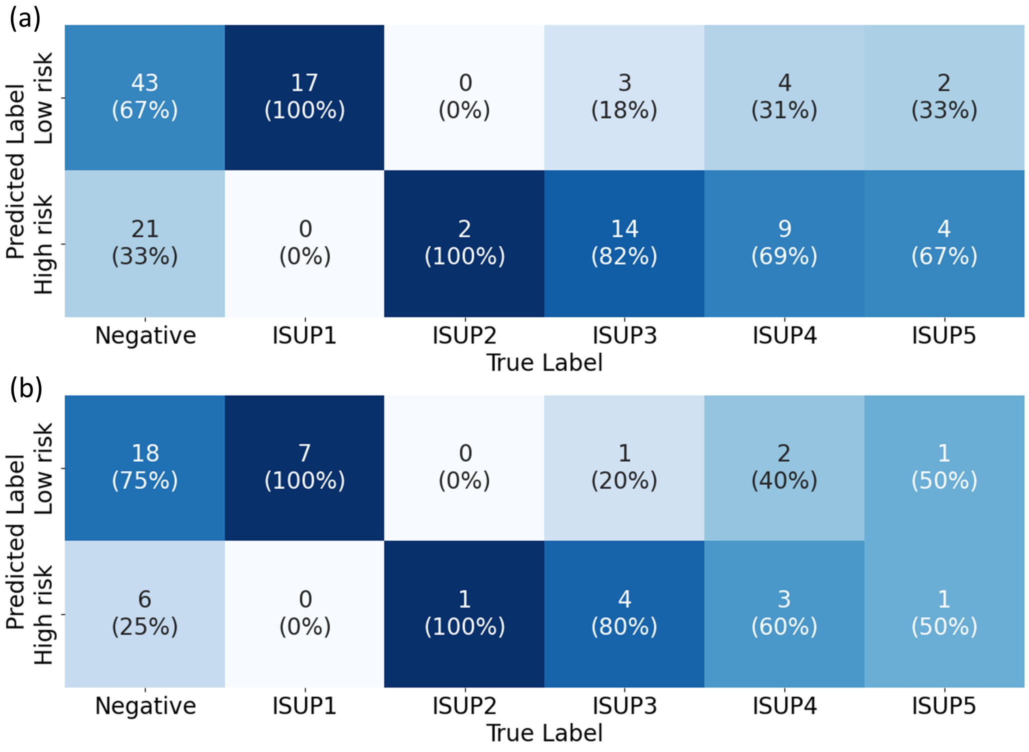


Figure B.1: Model performance on the test set. Contingency matrix (a) per slide and (b) per patient separating the true label by ISUP grade.

### Appendix C. Representation space for the whole cohort

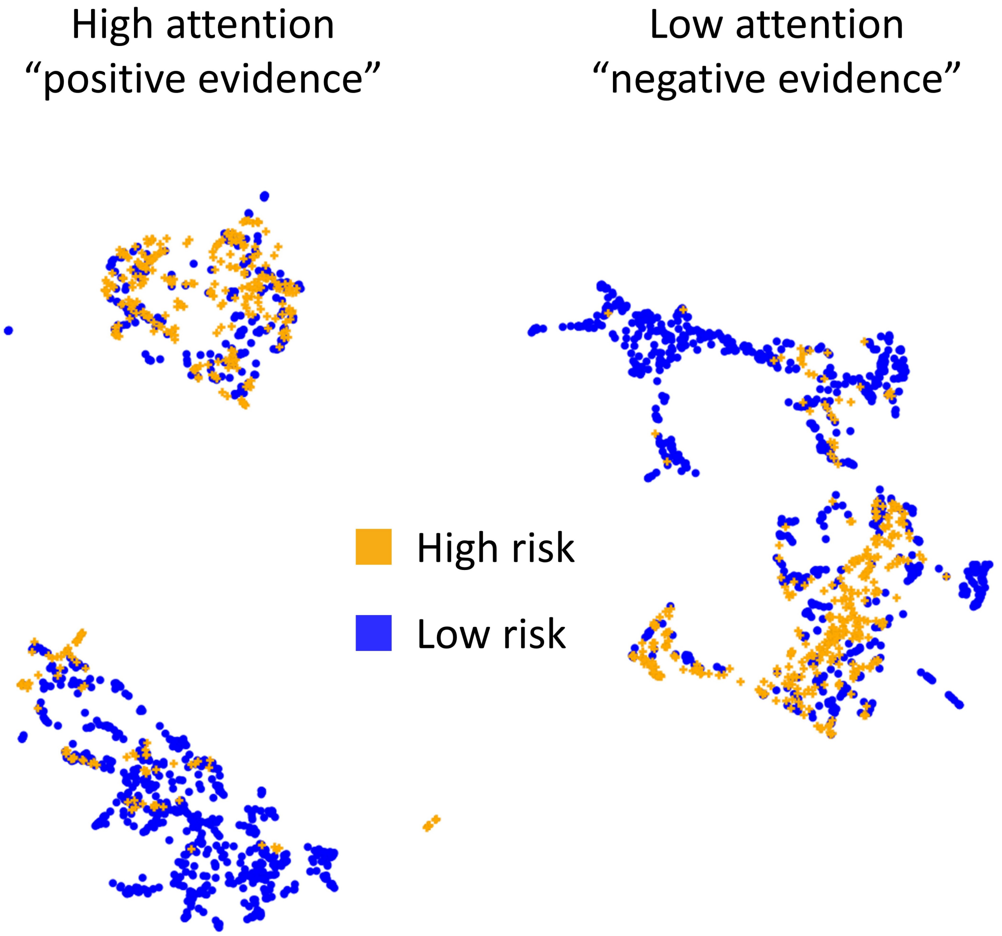


Figure C.2: UMAP space of the 10 most and least attended tiles from the complete test set.
